# Relationship between gait parameters and cognitive indexes in elderly adults

**DOI:** 10.1101/2022.09.23.22280301

**Authors:** Tania Aznielle-Rodríguez, Lídice Galán-García, Marlis Ontivero-Ortega, Karen Aguilar-Mateu, Ana M. Castro-Laguardia, Ana Fernández-Nin, Daysi García-Agustín, Mitchell Valdés-Sosa

## Abstract

**Purposes:** To select the summary cognitive index that is best predicted from spatio-temporal gait features (STGF) extracted from gait patterns.

**Methods:** 125 participants were recruited, including 40 young and 85 elderly adults. Their performances in different cognitive domains were evaluated through 12 cognitive measures obtained from five neuropsychological tests. Four summary cognitive indexes were calculated in each case: 1) the z-score of Mini-Mental State Examination (MMSE) from a population norm (MMSE z-score); 2) the sum of the absolute z-scores of the patient’s neuropsychological measures from a population norm (ZSum); 3) the patient’s scores for the first principal component (PC) of the set of neuropsychological test scores (PCCog); and 4) the Mahalanobis distance of each patient’s score from a population norm (MDCog). The gait patterns were recorded while they executed four walking tasks (normal, fast, easy- and hard-dual tasks). Sixteen STGF were measured using a body-fixed Inertial Measurement Unit. Dual-task costs were computed. The proportion of variances (R^2^) that PCA-STGF scores accounted for the four summary cognitive indexes and for the 12 cognitive variables across individuals were measured in multiple linear regressions. The confidence intervals for each R^2^ were estimated by bootstrapping the regression 1000 times.

**Results:** The mean values of R^2^ for the summary cognitive indexes were: 0.0831 for MMSE z-score, 0.0624 for ZSum, 0.0614 for PCCog, and 0.4751 for MDCog. The mean values of R^2^ for the 12 cognitive values ranged between 0.0566 and 0.1211. The multivariate linear regression was only statistically significant for MDCog, with the highest value of the R^2^ estimation.

**Conclusions:** Individual cognitive variables and most of the summary cognitive indices showed a weak association with gait parameters. However, the MDCog index showed a stronger association, explaining about 50 % of the variance. This suggests that this index can be used to study the relationship between gait patterns and cognition.

## 1 Introduction

The use of wearable sensors, such as Inertial Measurement Units (IMU), makes possible the recording of gait patterns and the extraction of spatio-temporal gait features (STGF) even outside of specialized laboratories (Ancillao et al., 2018). This has facilitated studies of the relationship between gait patterns and cognitive impairment within the community, especially in relation to aging (Montero-Odasso et al., 2012; Cohen, Verghese and Zwerling, 2016; Demnitz et al., 2016). The overall finding is that gait quality deteriorates in direct relationship to aging (Mahlknecht et al., 2013), and even more so when there is cognitive impairment (Montero-Odasso *et al*., 2012); Cohen, Verghese and Zwerling, 2016; Demnitz *et al*., 2016). However, the results are not entirely consistent and are difficult to generalize to new samples of subjects (Demnitz *et al*., 2016). This could be due to several problems.

The first problem is which overall summary measure of cognitive status is the most useful. A summary measure is needed to generate a grading of the subject’s cognitive deterioration. There are clinical tools that offer a fast global evaluation of cognition -e.g. the Mini-Mental State Examination (MMSE) (Folstein, Folstein and McHugh, 1975) or the Montreal Cognitive Assessment (MoCA) (Nasreddine *et al*., 2005)-potentially producing a single score. However, these instruments are semi-quantitative and have been reported to have low specificity and sensitivity (Demnitz *et al*., 2016; MacAulay *et al*., 2017; Carnero-Pardo, 2014). Additionally, the MMSE focuses primarily on the domains of language and memory, without assessing other cognitive domains potentially important to gait quality, such as the processing speed (Demnitz *et al*., 2016).

More quantitative measures can be obtained with many neuropsychological tests. However, each test usually examines a narrow cognitive domain (Schretlen, Bobholz and Brandt, 1996; Rabin, Barr and Burton, 2005; Brandt, 1991; Shum, McFarland and Bain, 1990). The idea of combining several cognitive variables into a single measure has been previously explored (Stijntjes *et al*., 2015; MacAulay *et al*., 2017; Byun *et al*., 2018; Beauchet *et al*., 2013), but we are left with a collection of (sometimes conflicting) measurements.

A second problem is that few studies have used normative data to correct for changes in cognitive test scores due to age, which is needed to assess the real severity of cognitive impairments (see Byun *et al*., 2018 for an exception). This means that the degree of the deviation of a participant’s cognitive functions from the population mean is generally not reported in studies of gait patterns. Thirdly, many studies employ a low number of participants, which may impair the reproducibility of the findings due to the low statistical power used (MacAulay *et al*., 2017; Montero-Odasso *et al*., 2009; Beauchet *et al*., 2017).

The present study was designed to try to overcome the above difficulties. We measured 12 cognitive variables obtained from five neuropsychological tests. To address the first problem, we computed four summary indexes from the battery of neuropsychological tests. The selected indexes reduce the dimensionality of the cognitive measures but retain quantitative and specific assessments of different cognitive domains. The indexes were the following: 1) the z-score of the MMSE, calculated from normative data, which has been used in different studies with older adults albeit not related to gait (Kondo *et al*., 2020; Janke *et al*., 2001) (MMSE z-score); 2) the sum of the z-scores absolute values of the deviation of the neuropsychological measures from normative data, previously used in other studies (Mielke *et al*., 2013) (ZSum); 3) the scores for the first principal component (PC) of the 12 cognitive variables (PCCog); and 4) the Mahalanobis distance of each patient’s score from a population norm (MDCog).

Principal Components Analysis (PCA) is frequently used for dimensionality reduction (Holtzer, Wang and Verghese, 2012). Moreover, the first PC of a set of psychological tasks is often used to assess Spearman’s G factor of (fluid) intelligence (Duncan, Burgess and Emslie, 1995).

On the other hand, the Mahalanobis distance has been previously used as a statistical measure of the deviation of a subject’s cognitive profile from the population mean profile (Elfadaly, Garthwaite and Crawford, 2016). It has been also used in clinical settings, for example: in assessing the cognitive profile of participants who were recovering from a stroke relative to the non-stroke controls (Tehan et al., 2018). This approach allows us to retain the quantitative aspect of neuropsychological tests while combining them into the single index MDCog. It also incorporates the use of deviations from the normative data while taking into consideration the correlations between features (avoiding spurious inflation of the estimated distance).

Here we examined the strength of the association, using multiple linear regression, of the four summary cognitive measures with a set of STGF extracted from the gait patterns. We hypothesized that the largest explained variance would correspond to the best cognitive indicator. Our results indicate that MDCog has the strongest association with STGF and thus could be useful for studies of the aging.

## 2 Materials and Methods

### 2.1 Participants

Around 200 participants from different health institutions in Havana city and from the Cuban Center for Neuroscience were screened for inclusion in the study, described in detail in Aznielle-Rodríguez *et al*., 2022a. The inclusion criteria were the participant’s agreement, young adults aged between 20 and 40 years and older adults ages above 60 years, and a Katz Index of independence ≥ 4, as evidence of functional independence without the need for supervision or external help in performing basic activities of daily life (Shelkey and Wallace, 2000; Katz *et al*., 1963). The participants with an inability to walk, major neurological disorders, diseases of the musculoskeletal system, or severe cognitive impairment were excluded. All participants underwent a neurological physical examination, a questionnaire, and a cognitive assessment previously to execute the walking tasks. 125 participants completed all the requirements and participated in the study. The sample was divided into 40 young adults (mean age 27.65 ± 4.14, 50 % women) and 85 older adults (mean age: 73.25 ± 6.99, 62.3 % women). Written informed consent was obtained from the participants or caregivers, and the study was approved by the ethics committee of CNEURO due to its compliance with the Helsinki declaration.

### 2.2 Experiment

Participants covered 40 m (20 m in each direction) in an obstacle-free and flat environment, executing four walking tasks: 1) walking freely at a comfortable self-chosen speed (NormalW); 2) walking at a comfortable self-chosen speed while simultaneously counting their steps, an easy cognitive task (EasyD); 3) walking at a comfortable self-chosen speed while simultaneously counting backward from 100, a hard cognitive task (HardD); and 4) walking as fast as possible without running (FastW).

Their gait patterns were recorded using an IMU (Bitalino RIoT, Plux Wireless Biosignals, Portugal), attached firmly to a velcro band, and placed near the body’s center of mass, on the lower back at the L3 spinal level. Data was processed as it was explained in Aznielle-Rodríguez *et al*., 2022. After computing the Initial Contact and Final Contact events of the gait cycles, 16 STGF were calculated for each walking task: 1) step time (StpT); 2) step time variability or step time coefficient of variation (StpTCoV); 3) stride time (StrT); 4) stride time variability or stride time coefficient of variation (StrTCoV); 5) cadence (Cd); 6) root mean square amplitude of the vertical acceleration (RMS); 7) double support duration or double support time (DSD); 8) single support duration or single support time (SSD); 9) swing duration feed 1 (SwDurF1); 10) swing duration feed 2 (SwDurF2); 11) stance duration feed 1 (StDurF1); 12) stance duration feed 2 (StDurF2); 13) step duration feed 1 (StepDurF1); 14) step duration feed 2 (StepDurF2); 15) step length (StepLg); and 16) speed (GS). All STGF were expressed in seconds (s), except Cd (steps/min), RMS (g), StepLg (m) and GS (m/s). STGF were computed using algorithms described in the literature (Zijlstra, 2004; Del Din, Godfrey and Rochester, 2016; Montero-Odasso *et al*., 2011; Yang *et al*., 2012; Jarchi *et al*., 2018; Bugané *et al*., 2012; Zijlstra and Hof, 2003; Del Din *et al*., 2016).

Additional to the 64 STGF, dual-task costs (DTC) for the STGF in the two dual-tasks were also calculated (32 costs). These costs (expressed as percentages) were calculated using (Montero-Odasso et al., 2017):

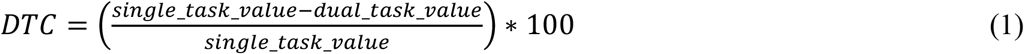

Finally, 96 measures (STGF and DTC) were obtained for each walking direction.

### 2.3 Cognitive assessment

The cognitive trait of the participants was assessed through five neuropsychological tests: 1) MMSE, as a global index of cognition (Folstein, Folstein and McHugh, 1975); 2) Attentional Span or Brief Test of Attention (BTA), as a measure of auditory divided attention (Schretlen, Bobholz and Brandt, 1996); 3) Trail Making Test (TMT), parts A and B, for assessing the attention, visuospatial abilities, mental flexibility, and executive functions (Rabin, Barr and Burton, 2005); 4) Hopkins Verbal Learning Test (HLVT) for memory evaluation, including immediate recognition and delayed recall (Brandt, 1991); and 5) Digit Symbol Substitution Test (DS), for the focused, selective and sustained attention as well as visual perception (Shum, McFarland and Bain, 1990). Twelve cognitive variables were extracted from the test results, which are shown in Table 1.

**Table 1.**
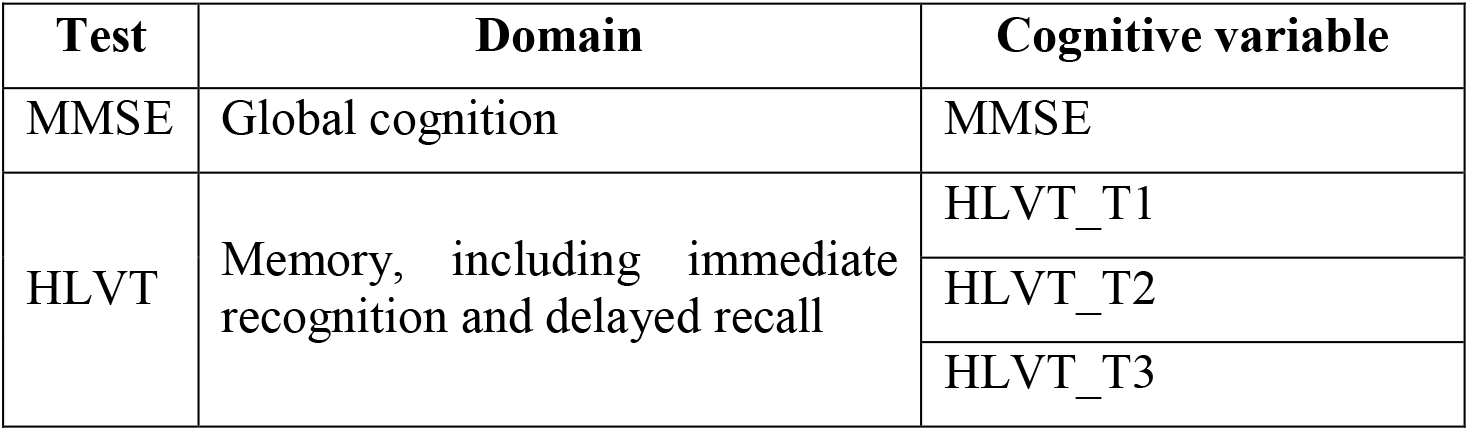
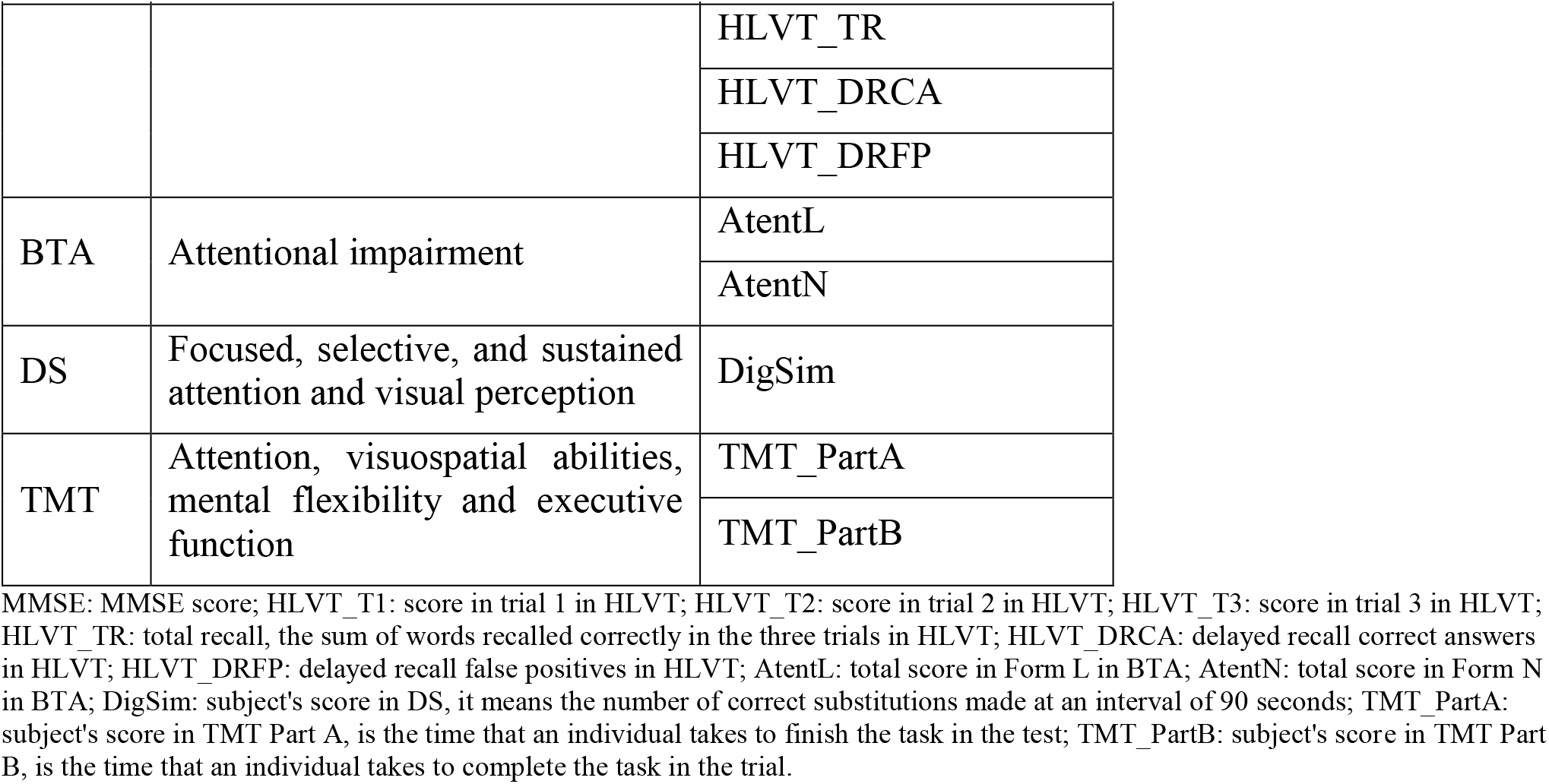
Cognitive variables extracted from the neuropsychological tests.

| Test | Domain | Cognitive variable |
| --- | --- | --- |
| MMSE | Global cognition | MMSE |
| HLVT | Memory, including immediate recognition and delayed recall | HLVT_T1 |
|  |  | HLVT_T2 |
|  |  | HLVT_T3 |

Relationship between gait parameters and cognitive indexes in elderly adults
| Test | Domain | Cognitive variable |
| --- | --- | --- |
|  |  | HLVT_TR |
|  |  | HLVT_DRCA |
|  |  | HLVT_DRFP |
| BTA | Attentional impairment | AtentL |
|  |  | AtentN |
| DS | Focused, selective, and sustained attention and visual perception | DigSim |
| TMT | Attention, visuospatial abilities, mental flexibility and executive function | TMT_PartA |
|  |  | TMT_PartB |

MMSE: MMSE score; HLVT\_T1: score in trial 1 in HLVT; HLVT\_T2: score in trial 2 in HLVT; HLVT\_T3: score in trial 3 in HLVT; HLVT\_TR: total recall, the sum of words recalled correctly in the three trials in HLVT; HLVT\_DRCA: delayed recall correct answers in HLVT; HLVT\_DRFP: delayed recall false positives in HLVT; AtentL: total score in Form L in BTA; AtentN: total score in Form N in BTA; DigSim: subject's score in DS, it means the number of correct substitutions made at an interval of 90 seconds; TMT\_PartA: subject's score in TMT Part A, is the time that an individual takes to finish the task in the test; TMT\_PartB: subject's score in TMT Part B, is the time that an individual takes to complete the task in the trial.

The same variables have normative data for the Cuban adult population obtained as part of an international collaborative study (Rivera *et al*., 2015; Arango-Lasprilla, Rivera, Aguayo, *et al*., 2015; Arango-Lasprilla, Rivera, Garza, *et al*., 2015; Arango-Lasprilla, Rivera, Rodríguez, *et al*., 2015).

### 2.4 Cognitive indexes

The four cognitive indexes were computed as follows:

1. MMSE z-score: The MMSE score is a value between 0 and 30, used in clinical practice as a global index of the cognitive trait. This score was transformed into the MMSE z-score using the Equation 2.

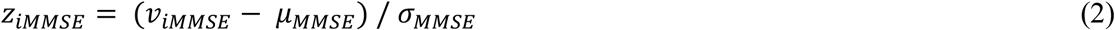

2. ZSum: This index was defined as the sum of the z-scores for the 12 cognitive variables, and was calculated using Equation 3.

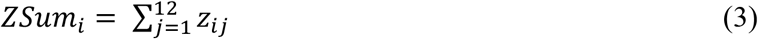

Where:

*z*_*ij*_ : is the z-score of the cognitive variable *j* for the subject *i*, calculated using the equation:

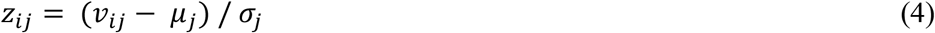

*µ*_*j*_ : mean value of the cognitive variable *j* in the population of normal subjects

*σ*_*j*_ : standard deviation of cognitive variable *j* in the population of normal subjects

These two population parameters were calculated using the normative regression functions of age and educational level-dependent mean values and standard deviations obtained from 306 normal subjects with an age ranging between 18 and 90 years, corresponding to the normative data for the Cuban adult population used in the collaborative study mentioned before (Rivera *et al*., 2015; Arango-Lasprilla, Rivera, Aguayo, *et al*., 2015; Arango-Lasprilla, Rivera, Garza, *et al*., 2015; Arango-Lasprilla, Rivera, Rodríguez, *et al*., 2015).

3. PCCog: PC was applied to the z-scores vector of the 12 cognitive variables, and the scores of the first component (explaining the largest proportion of variance) were retained.

4. MDCog: To measure the deviations from normative data, taking into consideration the correlations between the variables, this index was calculated using the Mahalanobis distance for each subject *i*, through the Equation 5.

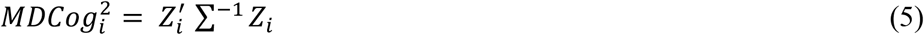

Where

*Z*_*i*_ : vector with the scores of the 12 cognitive variables for the subject *i*

∑^−1^ : represents the covariance matrix of the population norm.

### 2.5 Analysis

PCA was applied to create a new set of gait features (PCA-STGF) due to the high correlation between the 96 STGF. The components which explained more than 95 % of the variance were retained. The correlations between PCA-STG and the summary cognitive indexes were calculated. The proportion of variance (R^2^) that PCA-STGF scores could predict the values of summary cognitive indexes and the *z*_*j*_ scores of the 12 cognitive variables across individuals was measured in multiple linear regression. The confidence intervals for R^2^ were estimated by bootstrapping the regression 1000 times.

All data analysis was performed offline using MATLAB® (Mathworks Inc.).

## 3 Results

### Descriptive statistics of summary cognitive indexes

The histograms of the four summary cognitive indexes are presented in Figure 1.

**Figure 1.**
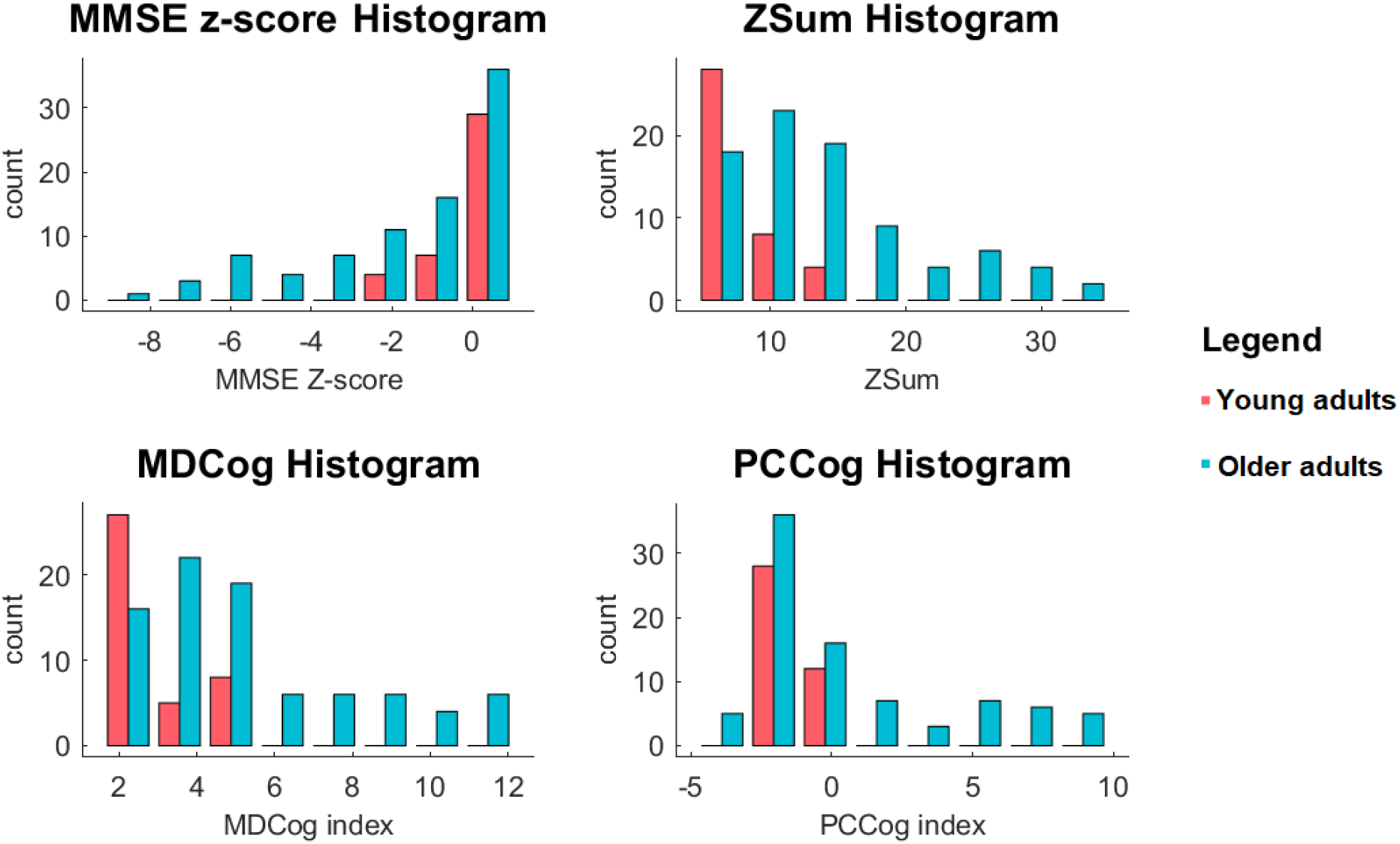
Histograms of the four summary cognitive indexes.

### Reducing dimensionality and redundancy in STGF

The new set of features PCA-STGF was obtained with orthogonal and independent components using a PCA. The first seven components explained over 95 % of the variance (highlighted with red dots in Figure 2), and were retained for further analyses. This result corroborated the high suspected redundancy between the STGF.

**Figure 2.**
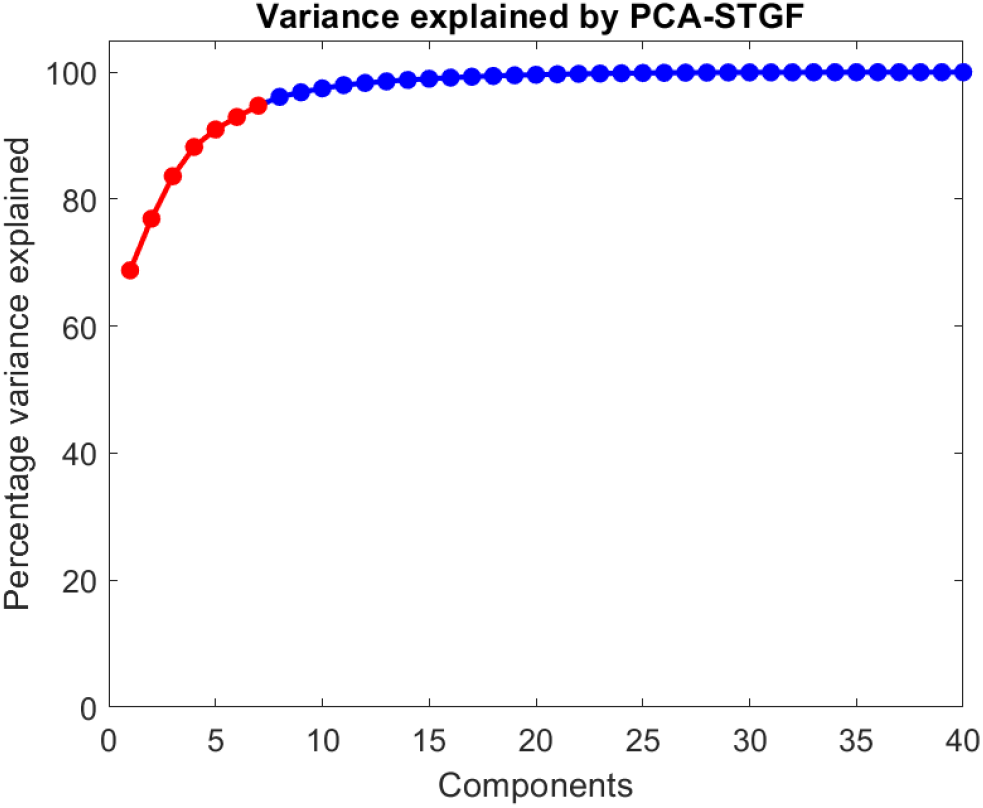
The cumulative sum of the variance accounted for the first 40 PCA-STGF components.

As shown in Figure 3, the contribution of each STGF to the first PCA component is similar in the four tasks. The features related to the description of the gait cycle had coefficients around 0.1, while Cd, RMS value, StepLg, and GS had coefficients around -0.1. The coefficients corresponding to gait variability features had values in the range between 0.07 and 0.1. The behavior of the features in the four tasks joined to the fact that the first PCA component explained the 68.79 % of the variance, suggesting that some walking conditions could be eliminated in the experiment without losing much information, thus shortening the total examination time.

**Figure 3.**
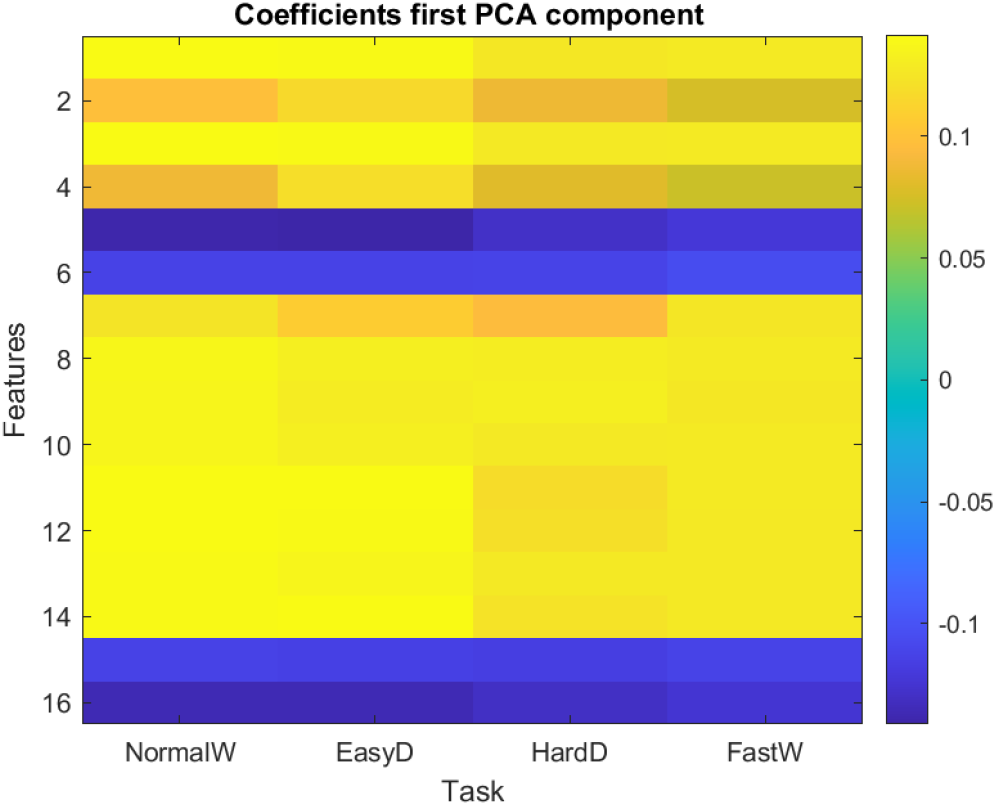
Contribution of the 16 STGF in each task to the first PCA component.

### PCA in cognitive variables

PCA was applied to the z-scores vector of the 12 cognitive variables. The cumulative sum of the new 12 components (PCA-CogVar) is shown in Figure 4. The first seven components explained over 95 % of the variance (highlighted in Figure 4 with red dots), and the first one explained the 40.98 % of variance. The score of the first component was retained as PCCog summary index.

**Figure 4.**
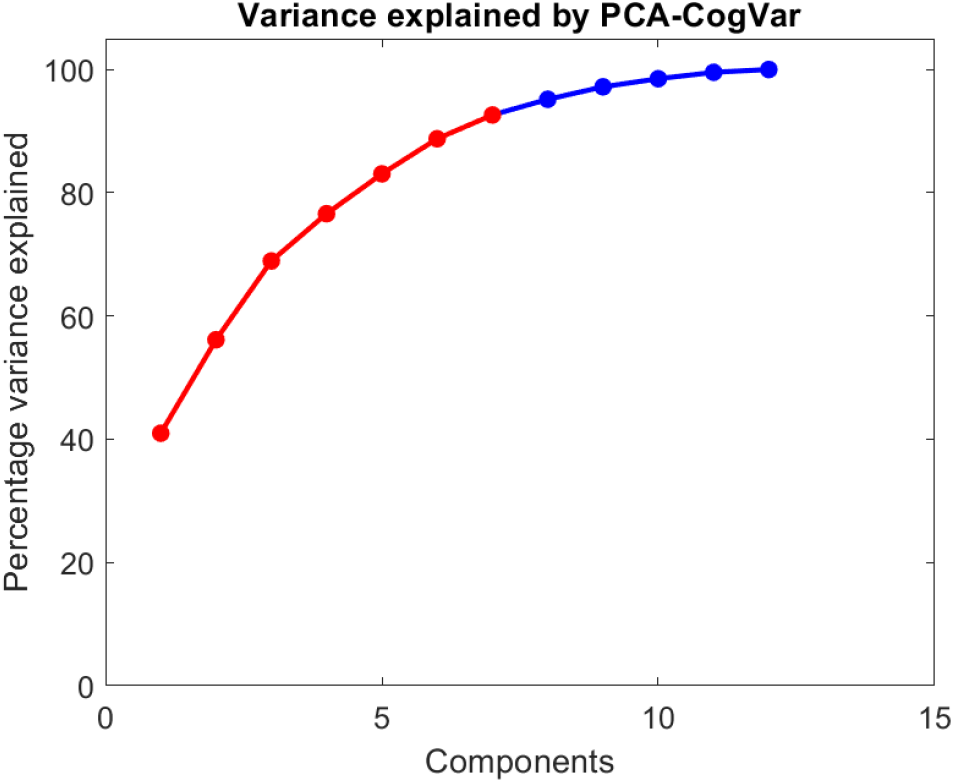
The cumulative sum of the variance accounted for the 12 PCA components.

### Relationship between cognitive variables and PCA-STGF

Multivariate linear regression with bootstrapping was applied to compare the predictability of the summary cognitive indexes using the first seven components of the PCA-STGF set. The results are shown in Table 2.

**Table 2.** Prediction of the summary cognitive indexes from PCA-STGF set.

| Variable | R<sup>2</sup> | F | p-Value | CI |
| --- | --- | --- | --- | --- |
| **Summary indexes** |  |  |  |  |
| MMSE z-score | 0.0831 | 1.5569 | 0.3098 | [0.0273-0.1625] |
| ZSum | 0.0624 | 1.1389 | 0.4592 | [0.0195-0.1328] |
| PCCog | 0.0614 | 1.1128 | 0.4524 | [0.0219-0.1102] |
| MDCog | 0.4751 | 15.7781 | 0.0000* | [0.3610-0.6028] |

\* Statistically significant

The multivariate linear regression was only statistically significant for MDCog, with the highest value of the R^2^ estimation. The confidence intervals for R^2^, estimated by bootstrapping the regression 1000 times, are also shown in the table. PCA-STGF reliably predicted much more of the variance for MDCog than the other summary cognitive indexes.

A new multivariate linear regression with bootstrapping was applied to compare the predictability of the best summary cognitive index (MDCog) regarding the z-scores of the individual cognitive variables, using the same set of gait variables. The results are shown in Table 3.

**Table 3.** Prediction of the z-scores of cognitive variables from PCA-STGF using a multivariate linear prediction with bootstrapping.

| Variable | R<sup>2</sup> | F | p-Value | CI |
| --- | --- | --- | --- | --- |
| HLVT_T1 | 0.0966 | 1.8222 | 0.1970 | [0.0408-0.1697] |
| HLVT_T2 | 0.1193 | 2.3133 | 0.1112 | [0.0519-0.2010] |
| HLVT_T3 | 0.1168 | 2.2541 | 0.1079 | [0.0572-0.1970] |
| HLVT_TR | 0.0901 | 1.6923 | 0.2422 | [0.0356-0.1663] |
| HLVT_DRCA | 0.0903 | 1.6966 | 0.2475 | [0.0323-0.1602] |
| HLVT_DRFP | 0.0566 | 1.0157 | 0.4862 | [0.0216-0.1033] |
| AtentL | 0.1031 | 1.9724 | 0.1903 | [0.0366-0.1874] |
| AtentN | 0.1211 | 2.3588 | 0.1141 | [0.0510-0.2108] |
| DigSim | 0.0872 | 1.6347 | 0.2689 | [0.0312-0.1619] |
| TMT_PartA | 0.0733 | 1.3457 | 0.3423 | [0.0302-0.1362] |
| TMT_PartB | 0.0683 | 1.2510 | 0.3982 | [0.0229-0.1302] |

Non-cognitive variable was statistically significant, as can be seen in Table 3.

## 4 Discussion

Here we explored four summary cognitive indexes to find which was best predicted by STGF. Since PCA revealed a significant redundancy between the STGF of different tasks, we used the seven components that explained 95 % of the variance. MDCog showed the closest relationship with gait features, since gait parameters explained more variance for MDCog than for the other summary indexes or any individual cognitive scores.

There are two possible explanations for this advantage of MDCog. First, combining indicators of ability in multiple areas into a global measure can help compensate for measurement noise in each test and provides an overall index of the subject’s cognitive function (Harvey, 2012). Second, it has been long known from factor analysis of multiple psychological tests that a general latent variable (Spearman’s G factor) explains a large proportion of the variance between individuals (Kane and Brand, 2003; Hoogendam *et al*., 2014). This G factor has been related to fluid intelligence (Duncan, Burgess and Emslie, 1995), effectiveness in executive function tasks (Crinella and Jen, 1999), and has been shown to decline with aging (Hoogendam *et al*., 2014). Our MDCog could use information related to this factor that taps essential processes needed to coordinate specific behaviors and manage multitasking. At the same time, it retains information about specific processes such as memory attention etc.

The lower performance of the other summary cognitive indexes could be due to several causes. Although the MMSE is a widely used global cognition index, its use has been criticized since its scores depend on the educational level of the subjects and on whether they are illiterate or not (Carnero-Pardo, 2014). Moreover, its sensitivity in discriminating MCI patients from those with normal cognitive aging is inadequate (MacAulay *et al*., 2017; Carnero-Pardo, 2014; Baek *et al*., 2016). On the other hand, the sum of the z-scores does not take into account the sign of the score of each variable or the correlation between them. Thus, a large pathological deviation on one variable will be added to a better than average performance, overestimating the real deterioration, and summing the z values of the highly correlated variables will also exaggerate scored anomaly.

Related to PCA, we used, as customary in the estimation of fluid intelligence, only the first component. In our case, it explained only 40.98 % of the variance in the cognitive variables, leaving 59.02 % unexplained. Thus, it discards much information, what is a disadvantage of this index compared to MDCog. This Spearman’s factor G by itself does not have a strong association with gait. MDCog takes into consideration the correlation between the results in each neuropsychological test and the distance of each subject from normative data. These factors, together with those stated above, probably explain the better results obtained with this index.

Many of the STGF used in this study describe the gait cycle, so there was a high correlation between them. This fact caused redundancy in the data, which has been pointed out by other authors as a problem in gait analysis (Olney *et al*., 1995). We used the same solution stated in many studies, which has been based on PCA (Olney *et al*., 1995; Mansour, Gorce and Rezzoug, 2017; Ren *et al*., 2017), for expecting that few components explained a reasonable amount of data. This statistical procedure converts the correlated characteristics into a smaller set of linearly uncorrelated components (Khera and Kumar, 2020). We found that the seven first PCA components explained most of the variance (95 %).

The limitations of this study are that the sample of older adults with cognitive impairment is smaller than the healthy one and both samples were not completely paired by sex and age. In future studies, it is recommended to delve into the relationship between the STGF domains and the cognitive domains, perhaps performing canonical correlation.

## Data Availability

All data produced in the present study are available upon reasonable request to the authors

## 5 Conclusions

The usefulness of MDCog as a summary and quantitative cognitive index status was proved. This new index is based on the Mahalanobis distance from each subject’s cognitive measure to the population norm and the interaction among these factors. MDCog can be used as an objective index instead the set of cognitive variables or other summary cognitive indexes explored in this study.

### Permission to reuse and Copyright

Copyright © 2022 Aznielle-Rodríguez, Galán-García, Ontivero-Ortega, Aguilar-Mateu, Castro-Laguardia, Fernández-Nin, García-Agustín, and Valdés-Sosa. This is an open-access article distributed under the terms of the Creative Commons Attribution License (CC BY). The use, distribution or reproduction in other forums is permitted, provided the original author(s) or licensor are credited and that the original publication in this journal is cited, in accordance with accepted academic practice. No use, distribution or reproduction is permitted which does not comply with these terms.

### Author Contributions

TA: gait recordings, data curation, software, analysis, and writing. LG: methodology, formal analysis, and reviewing. MO: methodology, formal analysis, reviewing, and editing. KA, AMC: methodology, cognitive assessment, AF, DG: methodology, gait recordings. MV: conceptualization, methodology, formal analysis, reviewing, and editing.

### Funding

This study was funded by the Flemish Interuniversity Council (VLIR-UOS) through the CU2017TEA436A103 project, awarded to Ghent University, Vrije Universiteit Brussel and Cuban Center for Neuroscience. Also, it was supported by the PN305LH13-050 project funded by the Oficina de Gestión de Fondos y Proyectos Internacionales, CITMA, Cuba.

## Acknowledgments

We thank the participants in this study, as well as the older adult’s caregivers who gave their consent. To Neisbet Blasco Fanego, Brenda Peón and José Aníbal Ojeda Núñez for the application and evaluation of the tests. To Jaime Menéndez Álvarez, Leisy Serrano Blanco and Gianna Arencibia for recording the gait patterns of the study participants.

